# Evaluating the environmental impact of clinical research: a life-cycle analysis of a French academic randomised clinical trial

**DOI:** 10.1101/2024.10.29.24316343

**Authors:** Claire Fougerou-Leurent, Louise Forteau, Maëlle Perrot-Loyer, Alain Renault, Catherine Mouchel, Chloe Rousseau, Enora Marion, Sabrina Cochennec, Anne Ganivet, Marie-Laure Gervais, Loïc Fin, Bruno Laviolle

**Affiliations:** Environnemental consulting Firm O2M Lab – Rennes, France; Pharmacology Department– Clinical Investigation Center Inserm1414 - Rennes University Hospital, France; Department of clinical research, Rennes University Hospital, France

**Keywords:** Clinical research, climate footprint, life-cycle analysis, eco-design

## Abstract

**Objectives:** To assess the environmental impact of an academic clinical trial, by adapting the life-cycle analysis methodology to clinical research.

**Design:** A retrospective, simplified complete life-cycle analysis (LCA), according to the EF 3.0 methodology. LCA is a standardised (ISO14040/44) method for assessing the environmental impacts of a product over its entire life cycle on several environmental issues. It is the balance of inputs and releases associated with the process, from conception to end-of-life.

**Setting:** A prospective, double-blind, randomised controlled trial in neurosurgery. The trial included 202 patients in 18 university hospitals across France.

**Participants:** Not applicable

**Intervention:** Not applicable

**Main outcome measures:** Fourteen impact indicators that could be combined into a single score, for the identification of hotspots of interest.

**Results:** Climate change (or greenhouse gas emissions), was the most important indicator, accounting for almost 30% of the single score. Greenhouse gas emissions were estimated at 31.6 tons of carbon dioxide equivalent. This was followed by the depletion of abiotic resources - fossil fuels (24%), resource use - minerals and metals (12%), and particulate matter emissions (8%). The main hotspots identified were patient’ transport and clinical research assistant travel for source data verification.

**Conclusions:** By using complete LCA approach, our study confirms that conducting a clinical trial has a significant environmental impact, particularly on climate change. The main identified hotspots were related to the transport of patients and clinical research assistants

**ARTICLE SUMMARY:** Strengths and limitations of this study

- This study describes the first complete life-cycle analysis of a clinical trial
- Fourteen environmental factors were assessed, appraising all trial activities, from conception to close-out
- This study complements those previously published in the process of establishing eco-design as a new paradigm for clinical research
- Only one clinical trial was assessed, limiting the transposability of the conclusions
- Due to the lack of published impact data for some activities, assumptions had to be made to estimate their environmental impact

## INTRODUCTION

Climate change is the leading global health threat of the 21st century. Modelling by the Intergovernmental Panel on Climate Change shows the possibility of average global warming of up to 5°C, which will pose an “existential threat” to humanity if current greenhouse gas emission trends are not curbed. In any case, the average temperature is expected to rise by 2°C by 2050 due to the carbon already emitted and the inertia of the economic system **Error! Reference source not found**.. The healthcare sector has a substantial environmental impact, contributing approximately 5% of global greenhouse gas emissions and generating significant waste, making it a major factor in the overall environmental footprint of modern societies [2]. In this context, the health system must anticipate two major changes: 1/ the evolution of the demand for care, linked to the impact of environmental changes on the health of the population, 2/ the transformation of the healthcare offer, linked to the double carbon constraint, i.e. reducing the carbon intensity of healthcare activity and finding substitutes for non-renewable resources.

While clinical trials are essential for identifying effective and safe treatments and preventing disease, they also have a significant environmental impact. In 2021, Adshead et al [3] estimated the greenhouse gas emissions of the 350,000 clinical trials registered on ClinicalTrials.gov to be 27.5 million tons of carbon dioxide equivalent (tCO2e), based on the results of a few pilot studies [4–6]. More recently, a small number of studies have examined carbon footprint in clinical trials, both in academic [7,8] and industrial [9,10] settings. Results for academically sponsored trials show that the total carbon footprint ranges from 15 to 765 tCO2e, and common hotspots have been identified as clinical trial unit (CTU) emissions, trial-specific patient assessments, and trial team meetings and travel.

Life Cycle Assessment (LCA) is a standardised method for assessing the potential environmental impacts of product systems, considering all processes and relevant environmental impacts associated with the life cycle of a product or service. The objective of our work was to adapt the LCA method to a phase 3 academic clinical trial in order to evaluate its environmental impacts. This work had two goals: to carry out a comprehensive assessment of the environmental impacts (not just carbon emissions) of an academic clinical trial, and to identify the main drivers of these impacts in order to discuss potential opportunities to mitigate the consequences of these hotspots.

## METHODS

### Life-cycle analysis

When we began our work, no guidance for evaluating the environmental impact of a clinical trial was publicly available. Therefore, a simplified attributional LCA was conducted from scratch using an iterative approach in accordance with ISO 14040/44 standards. The approach comprised the following four stages: 1/ Definition of goal and scope: life-cycle mapping; 2/ Inventory analysis: listing activities and stakeholders and collecting activity data; 3/ Impact assessment: calculating impact indicators and determining emission factors; 4/ Interpretation.

#### 1/ Definition of goal and scope: life-cycle mapping

We chose to assess the environmental impact of the SUCRE study (*Treatment of chronic subdural hematoma by corticosteroids: a prospective randomised study -* NCT02650609), sponsored by Rennes University Hospital (France). This study was selected because it was representative of a phase 3 French academic-sponsored clinical trial, terminated, and with available tracking data. This study was a prospective, double-blind, randomised controlled trial comparing methylprednisolone versus placebo in the treatment of chronic subdural hematoma without clinical and/or radiological signs of severity. Characteristics of the protocol have been published elsewhere [11]. The study included 202 patients in 18 university hospitals in France. The timeframe was nine years from funding application (2014) to submission for publication of the study results (2023).

In order to assess the life-cycle analysis of any product, process or service, the whole system needs to be established and understood. To do this, a series of information-gathering meetings were held with the clinical trials project team. These allowed for life-cycle mapping of the clinical trial and definition of the LCA boundaries. The different process steps were defined as follows: study design; regulatory procedures; logistical set-up; conduct of the study; closing and archiving (see Figure 1).

**Figure 1:**
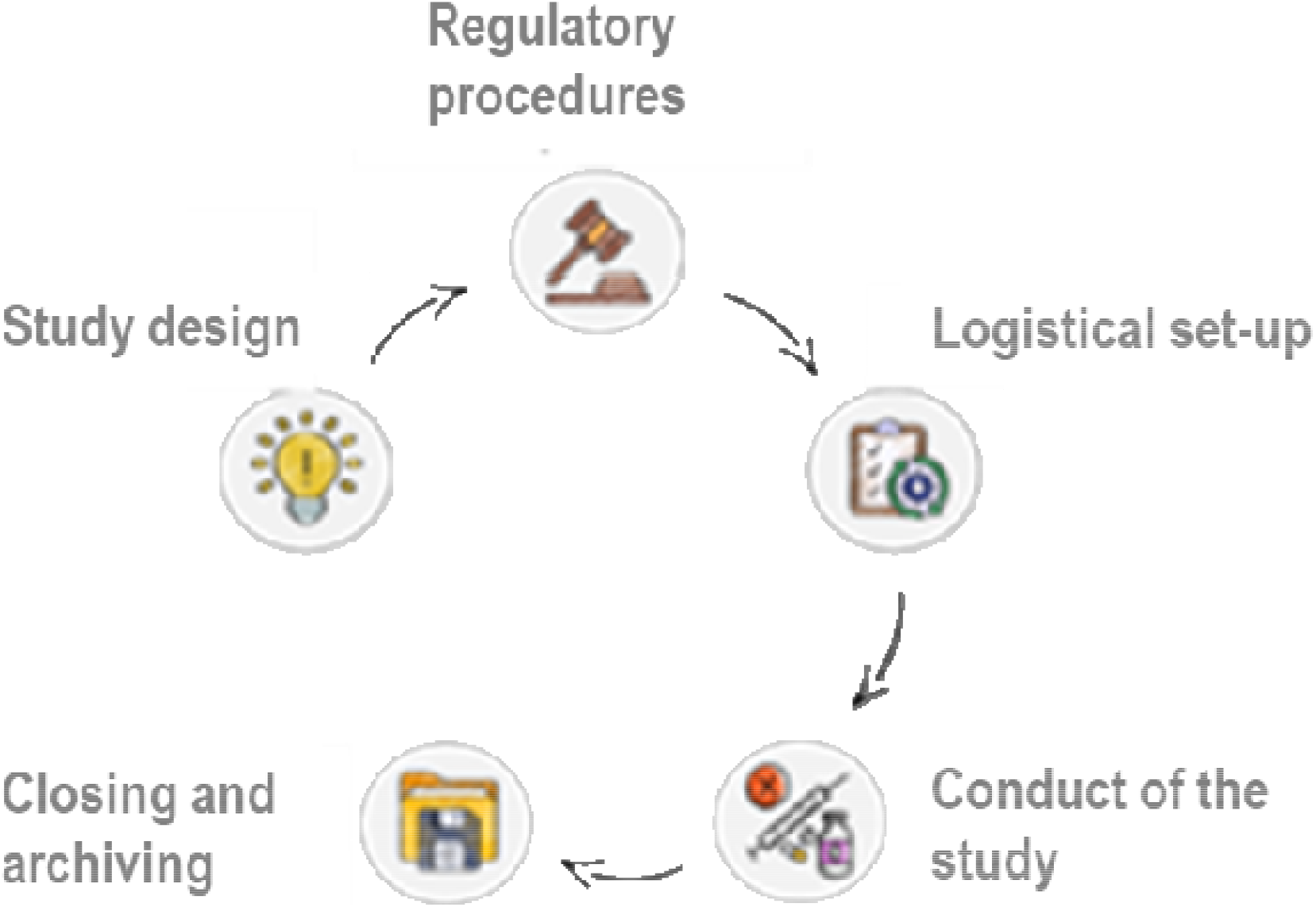
life-cycle of a clinical trial

#### 2/ Inventory analysis

The second step consisted of defining the core trial activities (see Table 1) and identifying the stakeholders. A data collection worksheet was produced so that activity data could be collected in a standardised way.

**Table 1:** Core trial activities included in the LCA. NB: For the following, the activity data from the investigating centres were extrapolated from the data collected for the Rennes centre, where appropriate in proportion to the number of patients included: patient transport, investigating team activity, number of CT scans performed.

| Core trial activity | Exclusions/Assumptions |
| --- | --- |
| Research personnel activity: utility consumption |  |
| Data collection and exchange (digital and paper): equipment (computers and screens, printers, paper), data storage, WIFI use, email exchanges, web requests, paper printing | Excluded: storage back-ups |
| Transport: on-site set-up, monitoring and close-out visits, patient transport to and from the clinical site, transport for presentation of results | Excluded: team commuting |
| Drug and placebo production and distribution | Assumption: proxy for drug composition and packaging weight |
| Consultations: utility consumption, medical devices and consumables | Excluded: electricity consumption in the consultation room, manufacture of CT scans, radiation emitted by CT scans |
| Waste (including drug waste) |  |

Identified stakeholders were asked to collect data related to their activities on the worksheet. Only activities undertaken for the clinical trial, over and above routine care, were included. Data were gathered from the trial documentation, Site Investigator File, sponsor’s trial specific databases and tracking files, mailboxes and electronic case report forms application. Global data that were not specific to the study came from two different databases: life cycle inventories (LCI) for processes such as sending an e-mail or performing an online search came from the Negaoctet database, LCI for energy mix and transportation (car, train, flight) came from the EcoInvent 3.9 database.

#### 3/ Impact assessment

A full LCA determines 16 indicators designed to quantify various environmental impacts associated with the process throughout its entire life cycle. These indicators can be categorised into several key areas (see Table 2) and grouped into a single score, calculated with weighting factors for each indicator. The weighting considers both the relative robustness of each indicator and the environmental challenges. The single score is expressed in Points (Pts), a unit based on the impact of an average European per year, in 2010. The lower the score, the lower the impact on the environment.

**Table 2:** indicators analysed in a full LCA.

|  |  |
| --- | --- |
| Climate Change | Measures the potential impact of greenhouse gases on global warming, usually expressed in CO<sub>2</sub> equivalents. |
| Resource Depletion | Fossil Fuel Depletion: assesses the consumption of non-renewable energy resources.<br>Mineral Resource Depletion: evaluates the depletion of mineral resources. |
| Ecosystem Quality | Acidification Potential: measures the potential for acid rain formation, which can harm ecosystems.<br>Eutrophication Potential: assesses the potential for nutrient enrichment in water bodies, leading to algal blooms and oxygen depletion.<br>Ecotoxicity: evaluates the potential toxic effects on ecosystems due to chemical emissions. |
| Human Health | Human Toxicity Potential: measures the potential health risks to humans from toxic substances.<br>Particulate Matter Formation: assesses the impact of particulate emissions on human respiratory health. |
| Water Use | Water Consumption: quantifies the total water used throughout the life cycle, considering both direct and indirect water use. |
| Land Use | Land Use Change: evaluates the impact of land use changes on biodiversity and ecosystem services. |
| Air Quality | Photochemical Ozone Creation Potential (POCP): measures the potential for ground-level ozone formation, which can cause respiratory problems and damage crops. |
| Additional Indicators | Ozone Depletion Potential (ODP): assesses the potential impact on the ozone layer.<br>Ionizing Radiation: evaluates the potential impact of radioactive emissions. |

The impact characterisation method used was EF 3.0 in SimaPro 3.5 software. By entering the LCIs of all the inputs and outputs from the SUCRE study, we were able to determine emission factors from 16 indicators and a single score for this clinical trial.

### Patient and public involvement

The LCA carried out in this work was a retrospective analysis of data collected during the SUCRE study. The data used were clinical trial documentation, activity data from the sponsor’s staff and interviews with trial staff and study site personnel. No patients participating in the trial were involved in the LCA and no personal information from the participants was collected or shared.

## RESULTS

The main indicator impacting the single score was climate change (28.5%), with total greenhouse gas (GHG) emissions estimated at 31,605 kgCO2e, or 156 kgCO2e per patient included (Table 3). This is roughly equivalent to the GHG emissions of three French citizens for one year, or nine return flights from Paris to New York.

**Table 3:** environmental impacts of the SUCRE study. The single score was estimated at 2.88 Pts.

| Impact category | Total | Contribution to the single score (%) |
| --- | --- | --- |
| Climate change | 31,605 kgCO<sub>2</sub>e | 28.5 |
| Depletion of abiotic resources - fossil fuels | 529,654 MJ | 23.5 |
| Resource use, minerals and metals | 0.3 kg Sb eq. | 12.2 |
| Particulate matter emissions | 1.6. 10<sup>-3</sup> disease incidence | 8.3 |
| Photochemical ozone formation | 139 kg NMVOC eq. | 5.7 |
| Acidification | 117 kg mol H<sup>+</sup> eq. | 4.5 |
| Ecotoxicity - freshwater | 248,071 CTUe | 3.9 |
| Ionising radiation | 7,397 kBq U235 eq. | 3.0 |
| Eutrophication - freshwater | 4.1 kg P eq. | 2.5 |
| Eutrophication - terrestrial | 340 Mol N eq. | 2.5 |
| Eutrophication - marine | 32.4 kg N eq. | 1.7 |
| Human toxicity, non-carcinogenic effects | 4.8.10<sup>-4</sup> CTUh | 1.3 |
| Human toxicity, carcinogenic effects | 2.4.10<sup>-5</sup> CTUh | 1.0 |
| Water use | 3,382 m<sup>3</sup> deprived | 0.9 |
| Land use | 153,596 Pt | 0.5 |
| Ozone depletion | $7 \cdot 10^{-4}$ kg CFC-11 eq. | 0.0 |

The other main impact factors were the depletion of fossil fuels (23.5%), the depletion of mineral resources (12.2%), and fine particle emissions (8.3%).

The “conduct of the study” phase was the main contributor to the environmental impact and this for all indicators analysed (between 95 and 100%). Within this phase, the major hotspots identified were patient transports for study consultations, clinical research assistants (CRA) transport, and study drugs and exams (see Figure 2).

**Figure 2:**
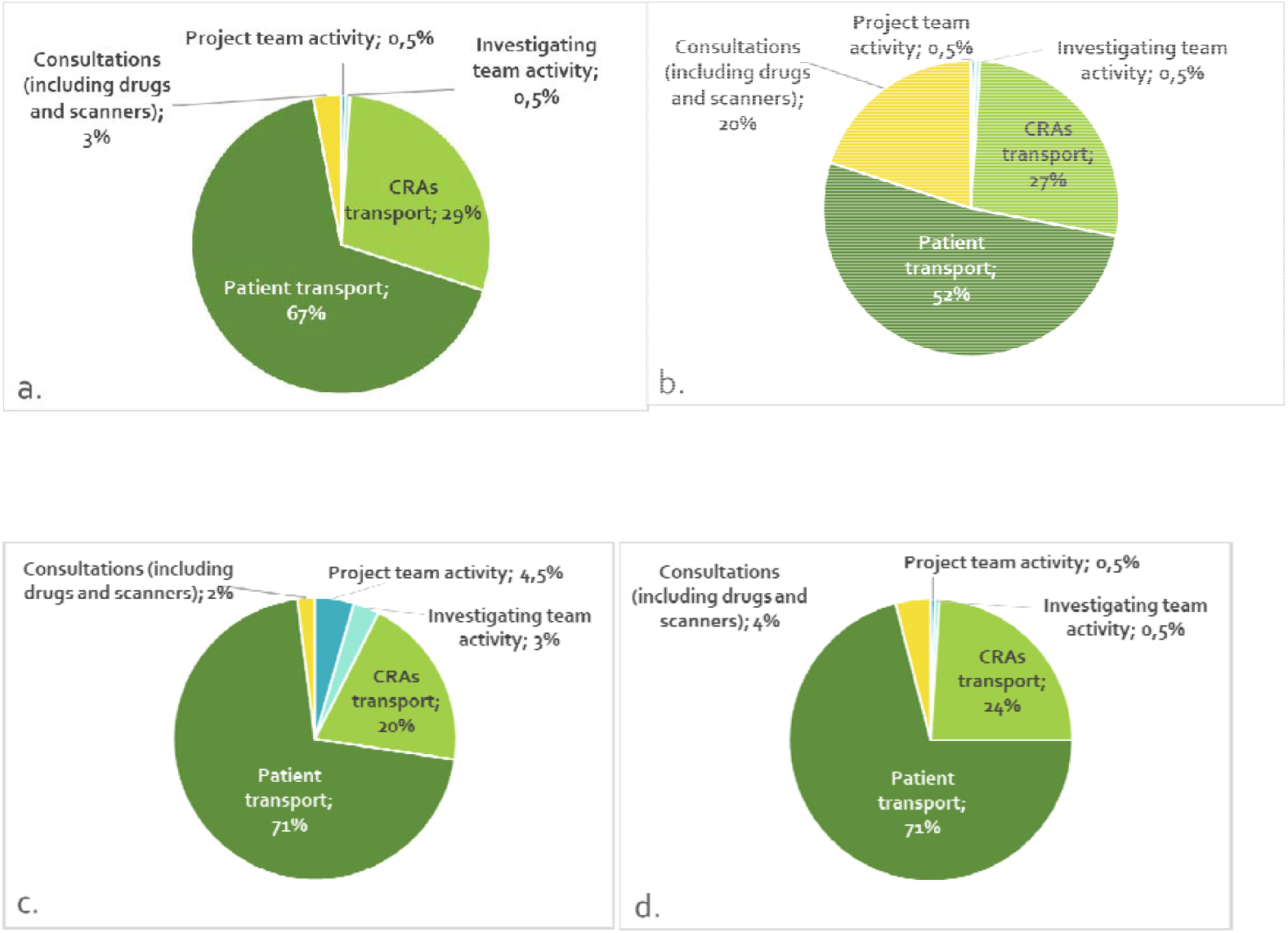
Impact of research activities on (a) climate change, (b) depletion of abiotic resources - fossil fuels, (c) resource use, minerals and metals, and (d) particulate matter emissions.

## DISCUSSION

In recent years, the issue of environmental sustainability in healthcare, including clinical research, has become increasingly important in light of the global climate threat. University hospitals are major actors in clinical research and, as well as implementing eco-design for care, they need to examine the sustainability of their practices in the context of clinical trials. To the best of our knowledge, our study is the first to report the results of a complete screening LCA carried out on a clinical trial. All previous studies only focused on GHG emissions from clinical trials using various methodologies, but none have questioned the justification for using this as a sole indicator. Our results show that climate change (due to GHG emissions) is a major contributor to the global environmental impact of a clinical trial. However, we also show that clinical trials have a significant impact on the depletion of fossil fuels, depletion of mineral resources and particulate matter emissions. Moreover, our data show that these impacts are related to the same activities i.e. transports of patients and team, allowing a concomitant reduction to be expected with the implementation of eco-design actions.

The carbon footprint of the SUCRE study was evaluated at 31.6 tCO2eq (corresponding to the climate change indicator), i.e. 156 kgCO2e per patient included. Although this result seems to be in line with those previously published [4–10], there are a number of methodological differences that make it difficult to compare studies. The methodology used to assess carbon emissions was initially based on the Greenhouse Gas Reporting Protocol [4–6], but more recent works have used a monocriteria life-cycle approach [9,10], or a methodology specifically adapted to clinical trials [7,8]. Harmonisation is necessary to standardise a specific methodology, such as that developed by Griffiths *et al* [7], and to adapt it to different contexts (academic/industrial, national), so that collaborative work can be envisaged.

Most previous studies reported CTU emissions, trial-specific patient assessments, meetings and travel by study personnel as major hotspots for carbon emissions [6–10]. Our results clearly confirm the main impact of trial-specific patient assessments, particularly due to patient transport, and trial staff transport; this major effect is found in the other three main indicators (depletion of fossil fuels, depletion of mineral resources and fine particle emissions). However, it should be noted that CTU emissions were not included in the hotspots of the SUCRE study, probably due to the scope of the assessment, as we chose not to include commuting. It seems to us that the subject of commuting is more a question of the sponsor’s organisation than of the management of the clinical trial. Moreover, as MacKillop *et al* previously highlighted [8], energy in France remains relatively low-carbon, which may contribute to the low share of CTU premises in carbon emissions.

## LIMITATIONS

The work presented here began in early 2023, in a context where there was no specific, recognised methodology for assessing the environmental impact of a clinical trial. We therefore opted for a generic LCA methodology, which we adapted to the ‘life cycle’ of a clinical trial. To ensure the feasibility of this first ‘screening’ LCA, we had to make a number of assumptions and exclude activities to be assessed (see § Methods). However, based on the results obtained, it is unlikely that the excluded data would have altered the respective contributions of the activities to the various indicators.

LCA is a relatively recent methodology, and the databases used for LCA analyses are frequently updated. Data on healthcare, pharmaceuticals, and web practices still require improvement. Additionally, access to some databases is restricted: EcoInvent and Negaoctet, for instance, are private, pay-per-use databases. HealthcareLCA, a database specific to healthcare, is one of the few open-access resources available, containing 7,000 emission factors for medical devices, drugs, medical interventions, companies and services. However, the calculation methodologies for these data are not standardised, and some do not fully adhere to LCA principles. Therefore, information from this database must be used cautiously, with careful consideration of the methodologies used to calculate the environmental impact.

The choice of the SUCRE study, an interventional drug study, limits its transposability to other study designs (particularly non-interventional studies). However, the choice of this study made it possible to evaluate activities that, *a priori*, had a significant environmental impact, such as the production and distribution of drugs and placebos, the monitoring of data from a multicentre trial, and performing examinations specific to the trial. Varying the types of studies assessed should now be a common objective, so that recommendations can be issued depending on the type of study and relevant good practice shared.

## PERSPECTIVES

Given the importance of travel in the environmental impact of clinical trials, certain practices that are developing internationally could provide solutions to reduce this impact, such as decentralised clinical trials and the use of remote monitoring. However, in addition to technical and organisational solutions, it is important to remember that the most effective way to reduce emissions is to reduce research waste, which results from research that is conducted but has no impact, for example because it lacks methodological rigour or unnecessarily duplicates a previous study [12,13]. Echoing Altman [14], we advocate that “less research, better research and research for the right reasons” also meets the goal of reducing the environmental impact of clinical research. It is in this spirit of minimisation that researchers should design and conduct clinical trials. To achieve this goal, the UK National Institute for Health and Care Research (NIHR) Carbon Reduction Guidelines highlight areas where good research design can reduce waste without compromising the validity and reliability of research [15]. However, it is also necessary to ask how stakeholders can adopt environmental management systems for clinical research and include the patient in this global consideration. Therefore, qualitative studies need to be conducted to understand the levers and barriers to the adoption of environmental approaches in clinical trials in order to establish a sustainable clinical research paradigm.

Finally, beyond the steps required at the level of sponsors and investigators, sustainability needs to be integrated at all levels of research, particularly in funding mechanisms [16] and research evaluation criteria [17, 18]. Eco-design of clinical research can only be achieved through a global paradigm shift.

## CONCLUSION

Through the first complete LCA of a clinical trial, our study has shown that a French academic clinical trial has a significant environmental impact and confirmed the interest of the use of GHG emissions as an indicator of this impact. Efforts must focus on the main hotspots identified in our study (namely patient transport and study staff travel) to reduce GHG emissions of clinical trials.

## Author statement

CFL and BL conceived of and presented the idea. LF contributed to study design. LF and CFL selected the clinical study from the Rennes University Hospital clinical trial portfolio for analysis. AR, CM, CR, EM, SC, AG, MLG and LF contributed to acquisition of data. MPL and LF created the data models for the analysis and contributed to the analysis and interpretation of data. CFL contributed to the data analysis and drafted the article. BL, AR and LF were involved in revising the article critically, and all authors approved the final version to be published.

### Patient and public involvement

Patients and/or the public were not involved in the design, or conduct, or reporting, or dissemination plans of this research.

### Patient consent for publication

Not applicable.

### Data availability statement

Data are available on request from the corresponding author.

